# Causal associations of education, lifestyle behaviors, and cardiometabolic traits with epigenetic age acceleration: a Mendelian randomization study

**DOI:** 10.1101/2022.07.12.22277528

**Authors:** Lijie Kong, Chaojie Ye, Yiying Wang, Jie Zheng, Shuangyuan Wang, Hong Lin, Zhiyun Zhao, Mian Li, Yu Xu, Jieli Lu, Yuhong Chen, Min Xu, Weiqing Wang, Guang Ning, Yufang Bi, Tiange Wang

**Affiliations:** Department of Endocrine and Metabolic Diseases, Shanghai Institute of Endocrine and Metabolic Diseases, Ruijin Hospital, Shanghai Jiao Tong University School of Medicine, Shanghai, China; Shanghai National Clinical Research Center for Metabolic Diseases, Key Laboratory for Endocrine and Metabolic Diseases of the National Health Commission of the PR China, Shanghai Key Laboratory for Endocrine Tumor, State Key Laboratory of Medical Genomics, Ruijin Hospital, Shanghai Jiao Tong University School of Medicine, Shanghai, China; MRC Integrative Epidemiology Unit (IEU), Bristol Medical School, University of Bristol, Oakfield House, Oakfield Grove, Bristol, BS8 2BN, United Kingdom

**Keywords:** Epigenetic ageing, Education, Lifestyle behaviors, Cardiometabolic traits, Mendelian randomization

## Abstract

**Background:** GrimAge acceleration (GrimAgeAccel) and PhenoAge acceleration (PhenoAgeAccel) are DNA methylation-based markers of accelerated biological ageing, standing out in predicting mortality and age-related cardiometabolic morbidities. Causal risk factors for GrimAgeAccel and PhenoAgeAccel are unclear.

**Objective:** To evaluate causal associations of 18 common modifiable socioeconomic, lifestyle, and cardiometabolic factors with GrimAgeAccel and PhenoAgeAccel.

**Methods:** We performed two-sample univariable and multivariable Mendelian randomization (MR), using summary-level data for GrimAgeAccel and PhenoAgeAccel derived from a genome-wide association study of 34,710 European participants. We used the inverse-variance weighted method as the main analysis, supplemented by three sensitivity analyses.

**Results:** Eleven and eight factors were causally associated with GrimAgeAccel and PhenoAgeAccel, respectively. Smoking initiation was the strongest risk factor (β [SE]: 1.299 [0.107] years) for GrimAgeAccel, followed by higher alcohol intake, higher waist circumference, daytime napping, higher body fat percentage, higher BMI, higher C-reactive protein, higher triglycerides, childhood obesity, and type 2 diabetes; whereas education was the strongest protective factor (β [SE] per 1-SD increase in years of schooling: -1.143 [0.121] years). Higher waist circumference (β [SE]: 0.850 [0.269] years) and education (β [SE]: -0.718 [0.151] years) were the leading causal risk and protective factors for PhenoAgeAccel, respectively. Sensitivity analyses strengthened the robustness of these causal associations, and the multivariable MR analyses demonstrated independent direct effects of the strongest risk and protective factors on GrimAgeAccel and PhenoAgeAccel, respectively.

**Conclusion:** Our findings provide novel quantitative evidence on modifiable causal risk factors for epigenetic ageing, and hint at underlying contributors and intervention targets to the ageing process.

**Key Points:**

- Genetically predicted higher educational attainment is the strongest protective factor for both GrimAgeAccel and PhenoAgeAccel, independent of causal lifestyle and cardiometabolic risk factors.
- Smoking is the leading causal lifestyle risk factor for GrimAgeAccel or PhenoAgeAccel, followed by alcohol intake and daytime napping. Adiposity traits are the leading causal cardiometabolic risk factors for GrimAgeAccel and PhenoAgeAccel, followed by type 2 diabetes, triglycerides, and C-reactive protein.
- This study provides novel quantitative evidence on modifiable causal risk factors for accelerated epigenetic ageing, indicating underlying contributors to the ageing process and promising intervention targets to promote healthy longevity.

## Introduction

Ageing involves the gradual accumulation of a decline in multiple biological functions over time, leading to increased risks of developing age-related diseases and mortality [1,2]. Although chronological ageing is uniform and unchangeable, the rate of biological ageing is variable and modifiable depending on individual genetics, environmental exposures, and health-related behaviors [3]. Of several potential types of biological age predictors (e.g., epigenetic clock, leukocyte telomere length, and transcriptomic predictors), the epigenetic clock that composed of DNA methylation at multiple cytosine-phosphate-guanine (CpG) sites is currently the best one, as it correlates well with age and predicts mortality across populations [4]. The epigenetic age acceleration is the difference between chronological age and epigenetic age, and represents accelerated biological ageing [5]. The second-generation epigenetic age acceleration indicators, namely GrimAge acceleration (GrimAgeAccel) and PhenoAge acceleration (PhenoAgeAccel), have been evolved to incorporate ageing-related traits, and stand out in terms of predicting mortality and age-related cardiometabolic morbidities [6-9].

Limited observational evidence has suggested that certain socioeconomic, lifestyle behaviors, and cardiometabolic traits may be related to GrimAgeAccel and PhenoAgeAccel [6,7,10,11]. Whether and to what extent modifiable risk factors influence GrimAgeAccel and PhenoAgeAccel, if causally established, could shed light on potential contributors to the ageing process and elucidate promising targets for preventing age-related diseases and improving healthy longevity [12,13]. Thus far, such causal evidence is scarce.

To fill this knowledge gap, we applied Mendelian randomization (MR) to evaluate the causal associations of 18 common modifiable socioeconomic, lifestyle and cardiometabolic factors with GrimAgeAccel and PhenoAgeAccel. The MR method uses genetic variants as instrumental variables (IVs) to infer causality among correlated traits. Since genetic variants are randomly allocated at conception, the MR study is less susceptible to confounding and reverse causality than conventional observational studies [14]. We conducted both univariable and multivariable MR analyses to discern if modifiable factors have independent direct causal effects on GrimAgeAccel and PhenoAgeAccel.

## Methods

### Study design

This MR study design is presented in **Figure 1**. We performed two-sample univariable and multivariable MR strictly following the STROBE-MR guidelines (**Supplementary Table 1**) [15]. To obtain unbiased estimates of the causal effects, the MR analysis should adhere to three fundamental assumptions [16]: first, the IVs are truly associated with the exposures; second, the IVs are independent of confounders of the relationship between exposures and outcomes (GrimAgeAccel and PhenoAgeAccel); third, the IVs influence the outcomes only through the exposures, but not any direct or indirect pathways. All data used in this MR study are publicly available. Ethical approval and informed consent had been obtained in all original studies.

**Figure 1.**
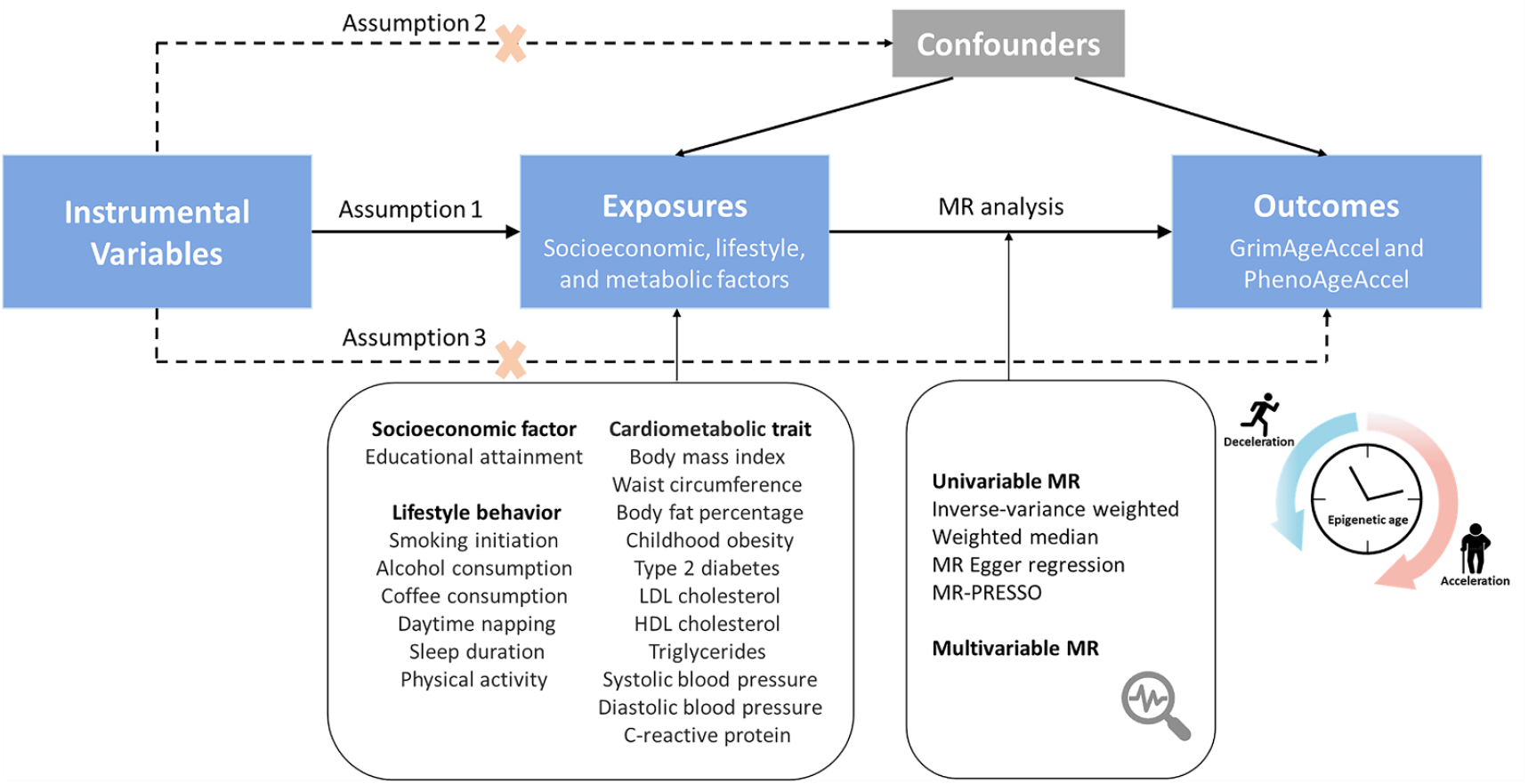
Study design and assumptions of the MR analysis. Assumption 1 indicates that the genetic variants proposed as instrumental variables should be robustly associated with the exposures; assumption 2 indicates that the used instrumental variables should not be associated with potential confounders of the relationship between exposures and outcomes; and assumption 3 indicates that the selected instrumental variables should influence the outcomes only through the exposures, not via alternative pathways. Abbreviations: GrimAgeAccel=epigenetic-age acceleration obtained using the GrimAge clock; HDL=high-density lipoprotein; LDL=low-density lipoprotein; MR=mendelian randomization; PhenoAgeAccel=epigenetic-age acceleration obtained using the PhenoAge clock; PRESSO=pleiotropy residual sum and outlier.

### Selection rationale and data sources of genetic instruments

We selected 18 common modifiable factors, including educational attainment, lifestyle behaviors (smoking initiation, alcohol intake, coffee consumption, daytime napping, sleep duration, and moderate-to vigorous physical activity [MVPA]), and cardiometabolic traits (body mass index [BMI], waist circumference, body fat percentage [BF%], childhood obesity, type 2 diabetes, low-density lipoprotein [LDL] cholesterol, high-density lipoprotein [HDL] cholesterol, triglycerides, systolic blood pressure [SBP], diastolic blood pressure [DBP], and C-reactive protein [CRP]). Definitions of the 18 modifiable factors are shown in **Supplementary Table 2**.

We extracted genetic variants for each of the modifiable factors from the largest available genome-wide association studies (GWASs) of European ancestry [17-29], ensuring minimum sample overlap with the genetic variants for GrimAgeAccel and PhenoAgeAccel (**Supplementary Table 3**). We included single nucleotide polymorphisms (SNPs) robustly associated with the 18 modifiable factors at the genome-wide significance (P <5×10-8). To select independent genetic variants, a stringent condition (linkage disequilibrium threshold of r^2^ <0.01) was set to minimize the influence of linkage disequilibrium which may bias the results of randomized allele allocation. Where SNPs for the exposures were not available in the GWAS summary statistics of GrimAgeAccel or PhenoAgeAccel, we used proxies of SNPs with r^2^ >0.8 as substitutes, by using the LDproxy search on the online platform LDlink (https://ldlink.nci.nih.gov/) [30].

### Data source for epigenetic age acceleration

The genetic associations with GrimAgeAccel and PhenoAgeAccel were extracted from a recent GWAS meta-analysis (summary statistics available at https://datashare.is.ed.ac.uk/handle/10283/3645), which included 34,710 European participants from 28 cohorts [31]. GrimAgeAccel and PhenoAgeAccel are two-generation epigenetic age acceleration indicators, expressing the biological ageing rate in years, of which GrimAgeAccel is more strongly associated with mortality than PhenoAgeAccel [11]. Detailed definitions of GrimAgeAccel and PhenoAgeAccel and data preparation in GWAS are shown in **Supplementary Table 2**.

### Statistical analyses

In the main analysis, we used the inverse-variance weighted (IVW) method to determine MR causal estimates (β coefficients with standard errors [SEs]) for associations of each modifiable factor with GrimAgeAccel and PhenoAgeAccel. The IVW combined the Wald ratio estimates of every single SNP in the set of IVs into one causal estimate using the random-effects meta-analysis approach [32]. To evaluate the robustness of the IVW estimates under different assumptions and to detect possible pleiotropy, we performed three sensitivity analyses, including the MR weighted median, the MR Egger, and the MR pleiotropy residual sum and outlier (MR-PRESSO) methods. The weighted median method selected the median MR estimate as the causal estimate, and provided a consistent causal estimate if over 50% of the weight in the analysis was derived from valid IVs [33]. The MR Egger method which allowed the intercept to be freely estimated as an indicator of pleiotropy was used to identify and adjust for the potential directional pleiotropic bias, but has limited precision [34]. The MR-PRESSO method was applied to detect and correct for any outlier SNP reflecting likely horizontal pleiotropic biases for MR causal estimates [35]. We evaluated the heterogeneity for the IVW estimates using the Cochran’s Q test [36], and identified the horizontal pleiotropy based on the p-value for the intercept in the MR-Egger model [34]. A false discovery rate (FDR) method was used to correct results for multiple testing, and FDR q-values were provided. In this study, strong causal evidence was defined as an association supported by the main analysis (FDR q-value <0.05) and at least one sensitivity analysis. Suggestive causal evidence was defined as a suggest association with P <0.05 and FDR q value ≥0.05 in the main analysis. Null causal evidence was defined as no statistically significant association revealed from the main analysis (P ≥0.05).

We further conducted multivariable MR (MVMR) analyses to assess whether the causal effects of the strongest protective factor and the strongest risk factor on GrimAgeAccel and PhenoAgeAccel were independent of other exposure factors [37]. Taking into account the effect size and significance of causal associations, we selected exposure factors with β coefficients >0.5 and P <0.05 in the main analysis as covariates in the adjustment models.

The two-sample MR analyses were conducted with the R packages ‘*TwoSampleMR’* and ‘*MRPRESSO*’ in R software (version 4.0.3). FDR q-values were estimated using the R package ‘*fdrtool*’.

## Results

### MR results for associations between 18 modifiable factors and GrimAgeAccel

Ten out of the 18 risk factors showed strong associations with increased GrimAgeAccel after FDR adjustment for multiple comparisons (**Table 1**). Smoking initiation was the strongest risk factor (β [SE]: 1.299 [0.107] years) for GrimAgeAccel, followed by higher alcohol intake (β [SE] per 1-SD increase: 0.899 [0.361] years), higher waist circumference (β [SE] per 1-SD increase: 0.815 [0.184] years), daytime napping (β [SE]: 0.805 [0.355] years), higher BF% (β [SE] per 1-SD increase: 0.748 [0.120] years), higher BMI (β [SE] per 1-SD increase: 0.592 [0.079] years), higher CRP (β [SE] per 1-SD increase: 0.345 [0.073] years), higher triglycerides (β [SE] per 1-SD increase: 0.249 [0.091] years), childhood obesity (β [SE]: 0.200 [0.075] years), and type 2 diabetes (β [SE]: 0.095 [0.041] years; **Figure 2a**). By contrast, educational attainment in years of schooling was the only one protective factor associated with decreased GrimAgeAccel (β [SE] per 1-SD increase: -1.143 [0.121] years). There was little evidence to support a causal association of other modifiable factors with GrimAgeAccel.

**Table 1.**
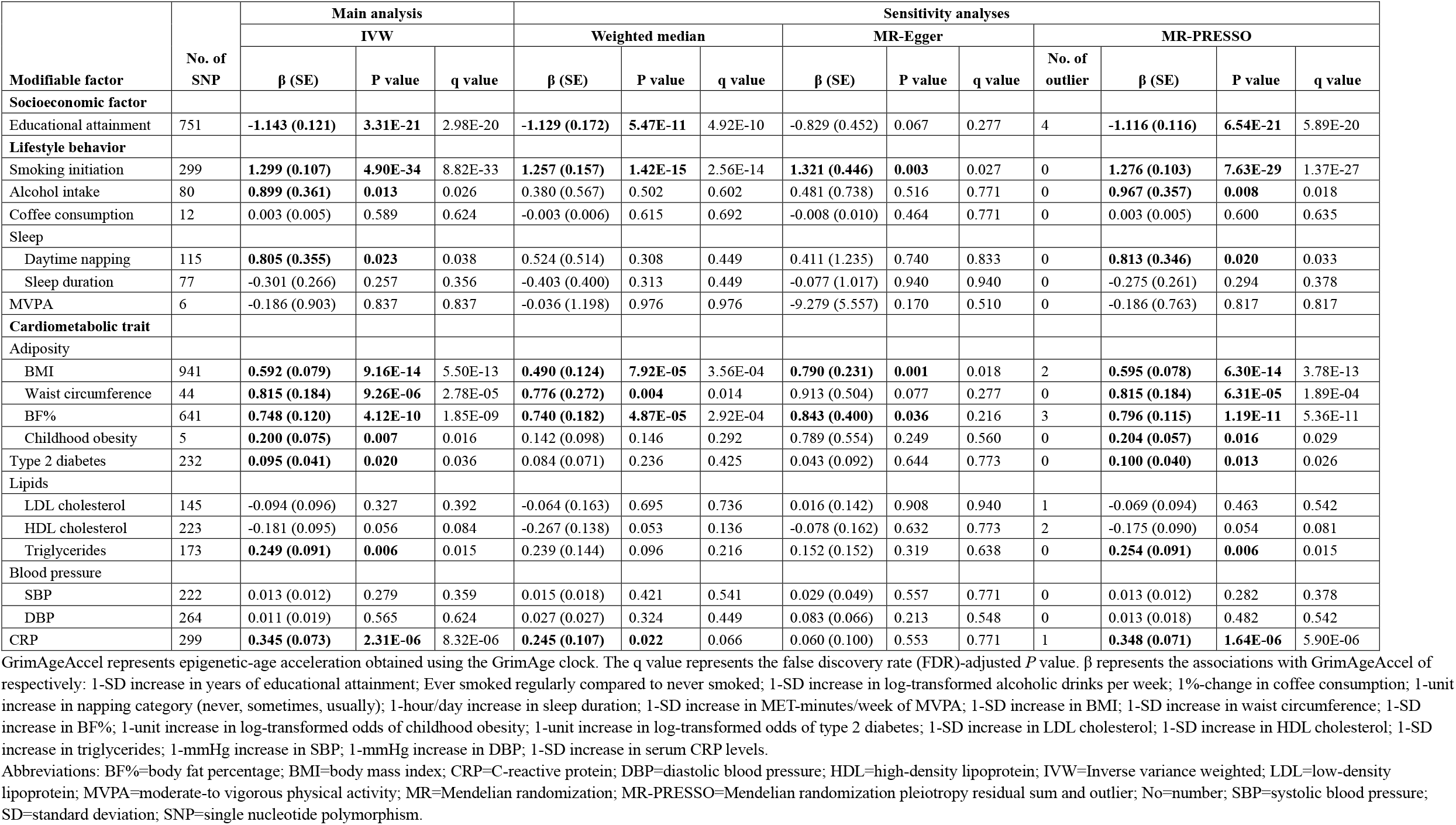
MR results of the associations between 18 modifiable factors and GrimAgeAccel.

| Modifiable factor | No. of SNP | Main analysis |  |  | Sensitivity analyses |  |  |  |  |  |  |  |  |  |
| --- | --- | --- | --- | --- | --- | --- | --- | --- | --- | --- | --- | --- | --- | --- |
|  |  | IVW |  |  | Weighted median |  |  | MR-Egger |  |  | MR-PRESSO |  |  |  |
|  |  | β (SE) | P value | q value | β (SE) | P value | q value | β (SE) | P value | q value | No. of outlier | β (SE) | P value | q value |
| Socioeconomic factor |  |  |  |  |  |  |  |  |  |  |  |  |  |  |
| Educational attainment | 751 | -1.143 (0.121) | 3.31E-21 | 2.98E-20 | -1.129 (0.172) | 5.47E-11 | 4.92E-10 | -0.829 (0.452) | 0.067 | 0.277 | 4 | -1.116 (0.116) | 6.54E-21 | 5.89E-20 |
| Lifestyle behavior |  |  |  |  |  |  |  |  |  |  |  |  |  |  |
| Smoking initiation | 299 | 1.299 (0.107) | 4.90E-34 | 8.82E-33 | 1.257 (0.157) | 1.42E-15 | 2.56E-14 | 1.321 (0.446) | 0.003 | 0.027 | 0 | 1.276 (0.103) | 7.63E-29 | 1.37E-27 |
| Alcohol intake | 80 | 0.899 (0.361) | 0.013 | 0.026 | 0.380 (0.567) | 0.502 | 0.602 | 0.481 (0.738) | 0.516 | 0.771 | 0 | 0.967 (0.357) | 0.008 | 0.018 |
| Coffee consumption | 12 | 0.003 (0.005) | 0.589 | 0.624 | -0.003 (0.006) | 0.615 | 0.692 | -0.008 (0.010) | 0.464 | 0.771 | 0 | 0.003 (0.005) | 0.600 | 0.635 |
| Sleep |  |  |  |  |  |  |  |  |  |  |  |  |  |  |
| Daytime napping | 115 | 0.805 (0.355) | 0.023 | 0.038 | 0.524 (0.514) | 0.308 | 0.449 | 0.411 (1.235) | 0.740 | 0.833 | 0 | 0.813 (0.346) | 0.020 | 0.033 |
| Sleep duration | 77 | -0.301 (0.266) | 0.257 | 0.356 | -0.403 (0.400) | 0.313 | 0.449 | -0.077 (1.017) | 0.940 | 0.940 | 0 | -0.275 (0.261) | 0.294 | 0.378 |
| MVPA | 6 | -0.186 (0.903) | 0.837 | 0.837 | -0.036 (1.198) | 0.976 | 0.976 | -9.279 (5.557) | 0.170 | 0.510 | 0 | -0.186 (0.763) | 0.817 | 0.817 |
| Cardiometabolic trait |  |  |  |  |  |  |  |  |  |  |  |  |  |  |
| Adiposity |  |  |  |  |  |  |  |  |  |  |  |  |  |  |
| BMI | 941 | 0.592 (0.079) | 9.16E-14 | 5.50E-13 | 0.490 (0.124) | 7.92E-05 | 3.56E-04 | 0.790 (0.231) | 0.001 | 0.018 | 2 | 0.595 (0.078) | 6.30E-14 | 3.78E-13 |
| Waist circumference | 44 | 0.815 (0.184) | 9.26E-06 | 2.78E-05 | 0.776 (0.272) | 0.004 | 0.014 | 0.913 (0.504) | 0.077 | 0.277 | 0 | 0.815 (0.184) | 6.31E-05 | 1.89E-04 |
| BF% | 641 | 0.748 (0.120) | 4.12E-10 | 1.85E-09 | 0.740 (0.182) | 4.87E-05 | 2.92E-04 | 0.843 (0.400) | 0.036 | 0.216 | 3 | 0.796 (0.115) | 1.19E-11 | 5.36E-11 |
| Childhood obesity | 5 | 0.200 (0.075) | 0.007 | 0.016 | 0.142 (0.098) | 0.146 | 0.292 | 0.789 (0.554) | 0.249 | 0.560 | 0 | 0.204 (0.057) | 0.016 | 0.029 |
| Type 2 diabetes | 232 | 0.095 (0.041) | 0.020 | 0.036 | 0.084 (0.071) | 0.236 | 0.425 | 0.043 (0.092) | 0.644 | 0.773 | 0 | 0.100 (0.040) | 0.013 | 0.026 |
| Lipids |  |  |  |  |  |  |  |  |  |  |  |  |  |  |
| LDL cholesterol | 145 | -0.094 (0.096) | 0.327 | 0.392 | -0.064 (0.163) | 0.695 | 0.736 | 0.016 (0.142) | 0.908 | 0.940 | 1 | -0.069 (0.094) | 0.463 | 0.542 |
| HDL cholesterol | 223 | -0.181 (0.095) | 0.056 | 0.084 | -0.267 (0.138) | 0.053 | 0.136 | -0.078 (0.162) | 0.632 | 0.773 | 2 | -0.175 (0.090) | 0.054 | 0.081 |
| Triglycerides | 173 | 0.249 (0.091) | 0.006 | 0.015 | 0.239 (0.144) | 0.096 | 0.216 | 0.152 (0.152) | 0.319 | 0.638 | 0 | 0.254 (0.091) | 0.006 | 0.015 |
| Blood pressure |  |  |  |  |  |  |  |  |  |  |  |  |  |  |
| SBP | 222 | 0.013 (0.012) | 0.279 | 0.359 | 0.015 (0.018) | 0.421 | 0.541 | 0.029 (0.049) | 0.557 | 0.771 | 0 | 0.013 (0.012) | 0.282 | 0.378 |
| DBP | 264 | 0.011 (0.019) | 0.565 | 0.624 | 0.027 (0.027) | 0.324 | 0.449 | 0.083 (0.066) | 0.213 | 0.548 | 0 | 0.013 (0.018) | 0.482 | 0.542 |
| CRP | 299 | 0.345 (0.073) | 2.31E-06 | 8.32E-06 | 0.245 (0.107) | 0.022 | 0.066 | 0.060 (0.100) | 0.553 | 0.771 | 1 | 0.348 (0.071) | 1.64E-06 | 5.90E-06 |

GrimAgeAccel represents epigenetic-age acceleration obtained using the GrimAge clock. The q value represents the false discovery rate (FDR)-adjusted *P* value. $\beta$ represents the associations with GrimAgeAccel of respectively: 1-SD increase in years of educational attainment; Ever smoked regularly compared to never smoked; 1-SD increase in log-transformed alcoholic drinks per week; 1%-change in coffee consumption; 1-unit increase in napping category (never, sometimes, usually); 1-hour/day increase in sleep duration; 1-SD increase in MET-minutes/week of MVPA; 1-SD increase in BMI; 1-SD increase in waist circumference; 1-SD increase in BF%; 1-unit increase in log-transformed odds of childhood obesity; 1-unit increase in log-transformed odds of type 2 diabetes; 1-SD increase in LDL cholesterol; 1-SD increase in HDL cholesterol; 1-SD increase in triglycerides; 1-mmHg increase in SBP; 1-mmHg increase in DBP; 1-SD increase in serum CRP levels.
Abbreviations: BF%=body fat percentage; BMI=body mass index; CRP=C-reactive protein; DBP=diastolic blood pressure; HDL=high-density lipoprotein; IVW=Inverse variance weighted; LDL=low-density lipoprotein; MVPA=moderate-to vigorous physical activity; MR=Mendelian randomization; MR-PRESSO=Mendelian randomization pleiotropy residual sum and outlier; No=number; SBP=systolic blood pressure; SD=standard deviation; SNP=single nucleotide polymorphism.

**Figure 2.**
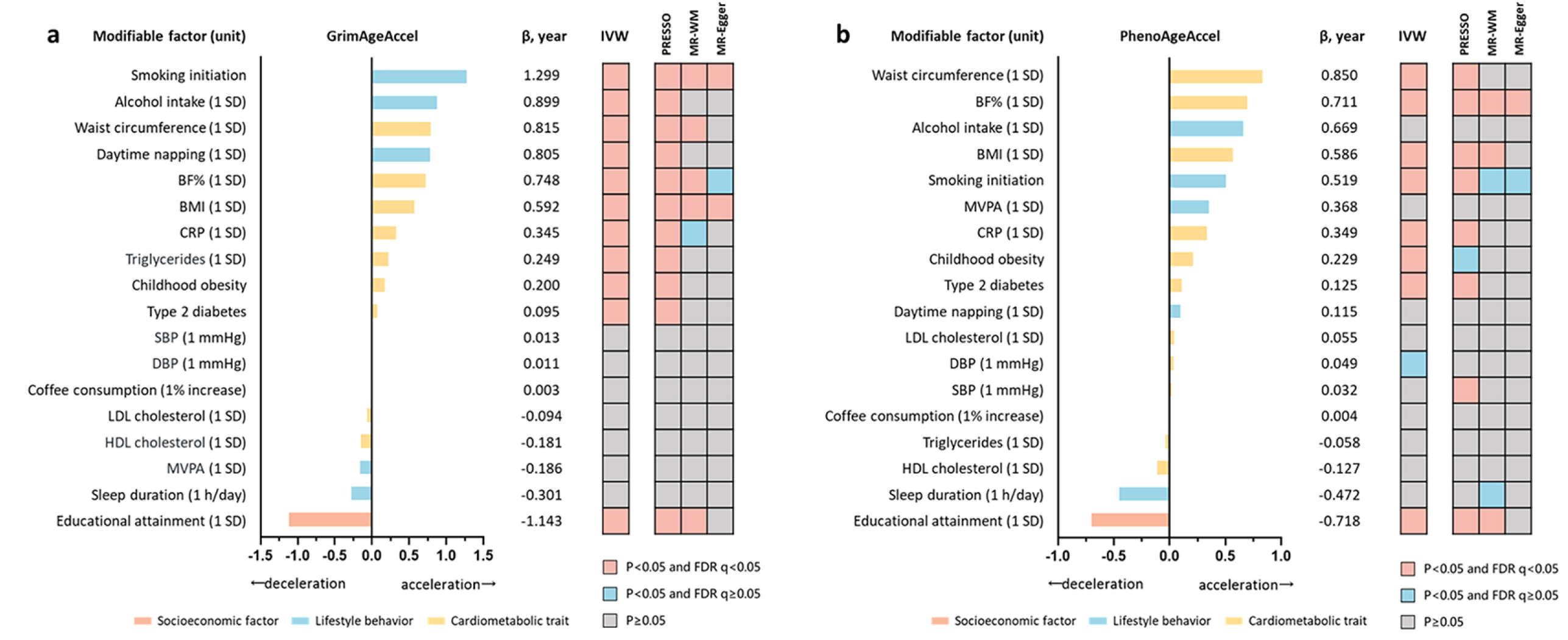
Overview of the findings on associations of 18 modifiable factors with GrimAgeAccel and PhenoAgeAccel. a. Causal associations between 18 modifiable factors and GrimAgeAccel; b. Causal associations between 18 modifiable factors and PhenoAgeAccel. GrimAgeAccel represents epigenetic-age acceleration obtained using the GrimAge clock; PhenoAgeAccel represents epigenetic-age acceleration obtained using the PhenoAge clock; β represents the effect of each modifiable factor on epigenetic-age acceleration. Red box indicates a strong association with p value <0.05 and FDR q value <0.05. Blue box indicates a suggestive association with p value <0.05 and FDR q value ≥0.05. Grey box indicates null association with p value ≥0.05. The IVW method was used for the main analysis. Sensitivity analyses included the MR-PRESSO, the MR-WM, and the MR-Egger methods. Abbreviations: BF%=body fat percentage; BMI=body mass index; CRP=C-reactive protein; DBP=diastolic blood pressure; HDL=high-density lipoprotein; IVW=inverse-variance weighted; LDL=low-density lipoprotein; MR=mendelian randomization; MVPA=moderate to vigorous physical activity; PRESSO=pleiotropy residual sum and outlier; SBP=systolic blood pressure; SD=standard deviation; WM=weighted median.

Associations of the above 11 modifiable factors with GrimAgeAccel were robust across sensitivity analyses with consistent effect directions and P <0.05 in at least one sensitivity analysis. The MR-Egger intercept tests indicated potential pleiotropy for CRP (P_intercept_ <0.05; **Supplementary Table 4**). Cochran’s Q-test showed possible heterogeneity for educational attainment, alcohol intake, BMI, BF%, type 2 diabetes, LDL cholesterol, HDL cholesterol, and CRP (P_h_ <0.05; **Supplementary Table 4**). However, with the exclusion of outlying SNPs, the MR-PRESSO analysis showed consistent results with the IVW analysis (**Table 1**).

### MR results for associations between 18 modifiable factors and PhenoAgeAccel

Eight modifiable factors showed strong associations with PhenoAgeAccel after FDR adjustment for multiple comparisons (**Table 2**). Higher waist circumference was the strongest risk factor (β [SE] per 1-SD increase: 0.850 [0.269] years) for PhenoAgeAccel, followed by higher BF% (β [SE] per 1-SD increase: 0.711 [0.152] years), higher BMI (β [SE] per 1-SD increase: 0.586 [0.102] years), smoking initiation (β [SE]: 0.519 [0.142] years), higher CRP (β [SE] per 1-SD increase: 0.349 [0.095] years), childhood obesity (β [SE]: 0.229 [0.095] years), and type 2 diabetes (β [SE]: 0.125 [0.051] years). Whereas the genetically predicted higher educational attainment in years of schooling was associated with decreased PhenoAgeAccel (β [SE] per 1-SD increase: -0.718 [0.151] years; **Figure 2b**). Suggestive causality was identified for the association between genetically predicted higher DBP and increased PhenoAgeAccel (β [SE] per 1-mmHg increase: 0.049 [0.024] years). No significant association was observed with PhenoAgeAccel for the other modifiable factors.

**Table 2.** MR results of the associations between 18 modifiable factors and PhenoAgeAccel.

| Modifiable factor | No. of SNP | Main analysis |  |  | Sensitivity analyses |  |  |  |  |  |  |  |  |  |
| --- | --- | --- | --- | --- | --- | --- | --- | --- | --- | --- | --- | --- | --- | --- |
|  |  | IVW |  |  | Weighted median |  |  | MR-Egger |  |  | MR-PRESSO |  |  |  |
|  |  | β (SE) | P value | q value | β (SE) | P value | q value | β (SE) | P value | q value | No. of outlier | β (SE) | P value | q value |
| Socioeconomic factor |  |  |  |  |  |  |  |  |  |  |  |  |  |  |
| Educational attainment | 751 | -0.718 (0.151) | 1.83E-06 | 1.65E-05 | -0.633 (0.225) | 0.005 | 0.030 | -0.361 (0.565) | 0.523 | 0.930 | 3 | -0.689 (0.145) | 2.52E-06 | 2.27E-05 |
| Lifestyle behavior |  |  |  |  |  |  |  |  |  |  |  |  |  |  |
| Smoking initiation | 299 | 0.519 (0.142) | 2.43E-04 | 0.001 | 0.404 (0.205) | 0.049 | 0.221 | 1.222 (0.589) | 0.039 | 0.240 | 1 | 0.545 (0.135) | 7.20E-05 | 3.24E-04 |
| Alcohol intake | 80 | 0.669 (0.441) | 0.130 | 0.213 | 1.134 (0.656) | 0.084 | 0.252 | 1.693 (0.923) | 0.070 | 0.252 | 0 | 0.737 (0.432) | 0.092 | 0.151 |
| Coffee consumption | 12 | 0.004 (0.008) | 0.600 | 0.735 | 0.002 (0.007) | 0.773 | 0.991 | -0.003 (0.015) | 0.861 | 0.930 | 0 | 0.004 (0.008) | 0.610 | 0.705 |
| Sleep |  |  |  |  |  |  |  |  |  |  |  |  |  |  |
| Daytime napping | 115 | 0.115 (0.500) | 0.818 | 0.818 | 0.014 (0.665) | 0.984 | 0.991 | 0.152 (1.742) | 0.931 | 0.931 | 0 | 0.083 (0.488) | 0.866 | 0.866 |
| Sleep duration | 77 | -0.472 (0.357) | 0.187 | 0.281 | -0.526 (0.462) | 0.255 | 0.417 | 2.771 (1.324) | 0.040 | 0.240 | 0 | -0.319 (0.357) | 0.375 | 0.519 |
| MVPA | 6 | 0.368 (1.138) | 0.747 | 0.791 | 0.262 (1.347) | 0.846 | 0.991 | -2.565 (6.947) | 0.731 | 0.930 | 0 | 0.368 (0.649) | 0.596 | 0.705 |
| Cardiometabolic trait |  |  |  |  |  |  |  |  |  |  |  |  |  |  |
| Adiposity |  |  |  |  |  |  |  |  |  |  |  |  |  |  |
| BMI | 941 | 0.586 (0.102) | 1.08E-08 | 1.94E-07 | 0.680 (0.170) | 6.33E-05 | 0.001 | 0.510 (0.298) | 0.087 | 0.261 | 4 | 0.605 (0.099) | 1.53E-09 | 2.75E-08 |
| Waist circumference | 44 | 0.850 (0.269) | 0.002 | 0.006 | 0.632 (0.339) | 0.062 | 0.223 | 0.739 (0.740) | 0.324 | 0.704 | 1 | 0.771 (0.249) | 0.003 | 0.009 |
| BF% | 641 | 0.711 (0.152) | 2.88E-06 | 1.73E-05 | 0.734 (0.227) | 0.001 | 0.009 | 1.666 (0.509) | 0.001 | 0.018 | 4 | 0.664 (0.145) | 5.75E-06 | 3.45E-05 |
| Childhood obesity | 5 | 0.229 (0.095) | 0.016 | 0.036 | 0.154 (0.123) | 0.211 | 0.380 | 0.140 (0.724) | 0.859 | 0.930 | 0 | 0.223 (0.071) | 0.025 | 0.050 |
| Type 2 diabetes | 232 | 0.125 (0.051) | 0.014 | 0.036 | 0.102 (0.080) | 0.205 | 0.380 | 0.056 (0.116) | 0.628 | 0.930 | 0 | 0.120 (0.049) | 0.016 | 0.036 |
| Lipids |  |  |  |  |  |  |  |  |  |  |  |  |  |  |
| LDL cholesterol | 145 | 0.055 (0.123) | 0.653 | 0.735 | -0.002 (0.174) | 0.991 | 0.991 | 0.169 (0.181) | 0.352 | 0.704 | 1 | 0.059 (0.123) | 0.633 | 0.705 |
| HDL cholesterol | 223 | -0.127 (0.109) | 0.243 | 0.336 | 0.068 (0.166) | 0.684 | 0.947 | 0.103 (0.187) | 0.583 | 0.930 | 0 | -0.121 (0.109) | 0.267 | 0.401 |
| Triglycerides | 173 | -0.058 (0.121) | 0.635 | 0.735 | 0.009 (0.196) | 0.964 | 0.991 | -0.202 (0.203) | 0.320 | 0.704 | 0 | -0.052 (0.121) | 0.666 | 0.705 |
| Blood pressure |  |  |  |  |  |  |  |  |  |  |  |  |  |  |
| SBP | 222 | 0.032 (0.016) | 0.053 | 0.095 | 0.033 (0.023) | 0.149 | 0.338 | -0.013 (0.065) | 0.840 | 0.930 | 1 | 0.038 (0.016) | 0.015 | 0.036 |
| DBP | 264 | 0.049 (0.024) | 0.037 | 0.074 | 0.048 (0.034) | 0.150 | 0.338 | 0.161 (0.084) | 0.055 | 0.248 | 0 | 0.042 (0.023) | 0.069 | 0.124 |
| CRP | 298 | 0.349 (0.095) | 2.49E-04 | 0.001 | 0.061 (0.150) | 0.684 | 0.947 | 0.020 (0.132) | 0.878 | 0.930 | 1 | 0.362 (0.093) | 1.16E-04 | 4.18E-04 |

PhenoAgeAccel represents epigenetic-age acceleration obtained using the PhenoAge clock; The q value represents the false discovery rate (FDR)-adjusted *P* value. $\beta$ represents the associations with PhenoAgeAccel of respectively: 1-SD increase in years of educational attainment; Ever smoked regularly compared to never smoked; 1-SD increase in log-transformed alcoholic drinks per week; 1%-change in coffee consumption; 1-unit increase in napping category (never, sometimes, usually); 1-hour/day increase in sleep duration; 1-SD increase in MET-minutes/week of MVPA; 1-SD increase in BMI; 1-SD increase in waist circumference; 1-SD increase in BF%; 1-unit increase in log-transformed odds of childhood obesity; 1-unit increase in log-transformed odds of type 2 diabetes; 1-SD increase in LDL cholesterol; 1-SD increase in HDL cholesterol; 1-SD increase in triglycerides; 1-mmHg increase in SBP; 1-mmHg increase in DBP; 1-SD increase in serum CRP levels.
Abbreviations: BF%=body fat percentage; BMI=body mass index; CRP=C-reactive protein; DBP=diastolic blood pressure; HDL=high-density lipoprotein; IVW=Inverse variance weighted; LDL=low-density lipoprotein; MVPA=moderate-to vigorous physical activity; MR=Mendelian randomization; MR-PRESSO=Mendelian randomization pleiotropy residual sum and outlier; No=number; SBP=systolic blood pressure; SD=standard deviation; SNP=single nucleotide polymorphism

Associations for the above eight modifiable factors were robust in sensitivity analyses with consistent effect directions and P <0.05 in at least one sensitivity analysis. There was no evidence of pleiotropy for these risk factors except for CRP (P_intercept_ <0.05; **Supplementary Table 5**). Potential heterogeneity was observed for educational attainment, smoking initiation, BMI, waist circumference, BF%, type 2 diabetes, and CRP (P_h_<0.05). One to four outliers were detected in the MR-PRESSO analyses; however, the associations remained consistent after removal of these outliers (**Table 2**). For SBP, with excluding the outlying SNP rs62523863, the MR-PRESSO analysis revealed a potentially positive association between genetically predicted higher SBP and PhenoAgeAccel (β [SE] per 1-mmHg increase: 0.038 [0.016] years).

### MVMR results for associations of the strongest protective and risk factors with GrimAgeAccel and PhenoAgeAccel

In MVMR analyses, the association between educational attainment (the strongest protective factor) and GrimAgeAccel remained significant with adjustment for smoking initiation, alcohol intake, waist circumference, daytime napping, BF%, or BMI (**Figure 3a**). Likewise, the significant association between smoking initiation (the strongest risk factor) and GrimAgeAccel persisted after adjustment for education, alcohol intake, waist circumference, daytime napping, BF%, or BMI. All covariates in MVMR analyses were selected based on their considerable effects on GrimAgeAccel in terms of size (β coefficients >0.5) and significance (P <0.05 in the main analysis).

**Figure 3.**
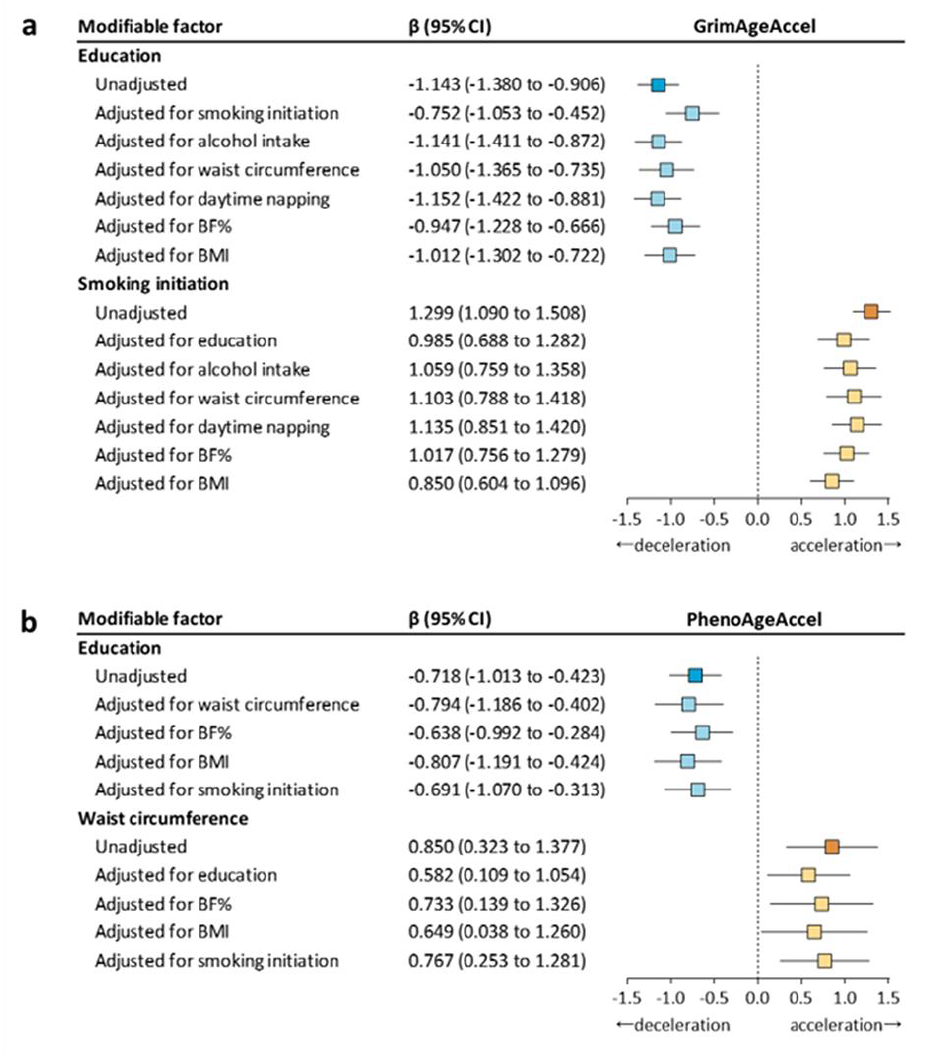
Multivariable MR assessing the effects of the strongest protective factor and risk factor on GrimAgeAccel and PhenoAgeAccel. a. Effect of the strongest protective factor and risk factor on GrimAgeAccel; b. Effect of the strongest protective factor and risk factor on PhenoAgeAccel. Causal estimates are β (95% CI) in years. Exposure factors with β coefficients >0.5 and P <0.05 in the main analysis as shown in Figure 2 were selected as covariates in the adjustment models. Abbreviations: BF%=body fat percentage; BMI=body mass index; CI=confidence intervals; MR=mendelian randomization.

The associations of education (the strongest protective factor) and waist circumference (the strongest risk factor) with PhenoAgeAccel remained significant with adjustment for waist circumference, BF%, BMI, or smoking initiation, which had considerable causal effects on PhenoAgeAccel (β coefficients >0.5; **Figure 3b**).

## Discussion

This MR study for the first time delineated potential causal relationships of 18 common modifiable factors with GrimAgeAccel and PhenoAgeAccel, the robust second-generation epigenetic age acceleration indicators. We identified strong evidence for 11 and eight factors associated with GrimAgeAccel and PhenoAgeAccel, respectively. Smoking initiation exhibited the greatest effect on increased GrimAgeAccel (1.299 years), followed by higher alcohol intake, higher waist circumference, daytime napping, higher BF%, higher BMI, higher CRP, higher triglycerides, childhood obesity, and type 2 diabetes; whereas educational attainment showed the greatest effect on decreased GrimAgeAccel (−1.143 years per 1-SD increase in years of schooling). Higher waist circumference and educational attainment were the leading causal risk and protective factors associated with PhenoAgeAccel, respectively; BF%, BMI, smoking initiation, CRP, childhood obesity, and type 2 diabetes were also associated with increased PhenoAgeAccel. Multiple sensitivity analyses further strengthened the robustness of these causal relationships, and the MVMR analyses demonstrated the independent direct effects of the strongest risk and protective factors on GrimAgeAccel and PhenoAgeAccel, respectively. Suggestive causality was identified for the association between higher DBP and increased PhenoAgeAccel.

In this study, educational attainment was the major protective factor for both GrimAgeAccel and PhenoAgeAccel, and this causal effect was largely independent of other causal factors, such as smoking and adiposity-related traits. Educational attainment is a strong proxy for socioeconomic status and a more upstream determinant of health, with broad implications for a person’s life-long lifestyle behaviors and health-promoting resources [38]. A recent study using UK Biobank data has documented that each 1-year increase in genetically determined educational attainment was associated with equivalently 4.2 years of age-related increases in telomere length [39]. Telomere length and epigenetic age acceleration metrics (e.g., GrimAgeAccel and PhenoAgeAccel) point towards distinct mechanisms of the ageing process that are marked by telomeres and the DNA methylation-based epigenetic clocks, both of which are independently associated with chronological age and mortality risk [40]. Our findings, together with those of the UK Biobank, highlight the important impact of educational attainment on biological ageing rates from two different aspects of the ageing process. Therefore, public health strategies aimed at reducing educational inequalities and improving educational attainment may slow the biological ageing rate and help reduce age-related health burdens.

This MR study identified several common lifestyle behaviors, including smoking initiation, alcohol intake, and daytime napping, causally associated with increased GrimAgeAccel or PhenoAgeAccel (smoking initiation only), which were consistent with previous evidence from observational studies [6,7]. Smoking methylation proxy is a component of the GrimAge clock [7], thus it is not surprising that smoking initiation exhibited a large effect on increased GrimAgeAccel in this study, and the MVMR analysis further confirmed that the effect of smoking on GrimAgeAccel was independent of other causal lifestyle behaviors and adiposity-related traits. Similarly, genetically determined smoking has been associated with shorter telomere length in the UK Biobank [41], and evidence from the Danish Health Interview Survey suggested that the life expectancy of a heavy smoker was a little more than seven years shorter than that of a never smoker [42]. Our study also provided strong MR evidence that genetically predicted higher alcohol intake was associated with increased GrimAgeAccel. A study composed of the Hannum cohort and Family and Community Health Studies cohort found that the relationship between alcohol use and the first-generation epigenetic clocks seemed to be nonlinear [42]. However, given the dose-dependent relationship of alcohol intake with all-cause mortality and cancers [43], our findings suggest that reducing alcohol intake is necessary to decrease GrimAgeAccel and retard the overall health loss. Previous observational studies also reported positive or inverse correlations of other lifestyle behaviors such as MVPA, coffee, and sleep duration with GrimAgeAccel, PhenoAgeAccel, or mortality [11,44,45]. Nevertheless, in this study, there was little evidence supporting these associations were causal. The discrepancy between our findings and previous observations may result from the potential confounding or reverse causation in conventional observational studies. Moreover, the non-linear association patterns, as in the case of sleep duration and mortality, might also partially explain the inconsistent results [46]. Therefore, our null findings should be cautiously interpreted.

Interestingly, of all cardiometabolic traits (i.e., adiposity traits, type 2 diabetes, triglycerides, and CRP) which causally increased GrimAgeAccel or PhenoAgeAccel, adiposity traits were the most dominant traits. Emerging observational studies have pronounced the positive associations of BMI with GrimAgeAccel and PhenoAgeAccel [11], and a meta-analysis of 87 observational studies showed each 5-kg/m^2^ higher BMI corresponded to about 1 year of age-related decrease in telomere length [46]. Our study further provided strong evidence for causal associations of various adiposity phenotypes, including waist circumference, BF%, BMI, and childhood obesity, with increased GrimAgeAccel and PhenoAgeAccel. In this study, type 2 diabetes, triglycerides, and CRP showed strong but modest effects (β coefficients ≤0.5) on GrimAgeAccel or PhenoAgeAccel, which was consistent with the findings of the UK Biobank study on telomere length [39].

This MR study yields new insights into modifiable causal factors, including education, lifestyle behaviors, and cardiometabolic traits, associated with accelerating or decelerating epigenetic ageing. This study brings us one step closer to understanding the potential contributors to the ageing process and provides promising intervention targets for healthy ageing. Given the enormous burden induced by age-related morbidity and mortality, strategies to reduce educational inequalities, promote healthy lifestyles primarily through reducing smoking, alcohol intake, and daytime napping, and improve cardiometabolic traits, specifically adiposity, type 2 diabetes, triglycerides, and CRP, to slow biological ageing rate are imminent.

In this study, we included independent and genome-wide significant SNPs as instruments for each of the modifiable factors to ensure the first MR assumption was fulfilled. Moreover, we applied strict criteria strengthened by the FDR-corrected significance and the cross-validations by main and sensitivity analyses to draw robust causal conclusions. However, several limitations merited consideration. First, we found potential pleiotropy from the MR-Egger intercept test for CRP. However, we conducted MR-PRESSO analysis, and the association remained consistent after removal of outlying SNPs. Second, we could not rule out the possibility that the associations of certain modifiable risk factors such as alcohol intake and sleep duration with GrimAgeAccel or PhenoAgeAccel may be non-linear. Future studies with individual-level data are warranted to confirm the linear or non-linear relationships. Third, to ensure the consistency of genetic background, this MR study was performed only in European-ancestry participants, thus the generalization of our results to other ethnic groups should be cautious.

## Conclusions

This MR study provided novel quantitative evidence on modifiable causal socioeconomic, lifestyle, and cardiometabolic factors for accelerated epigenetic ageing. Our findings shed light on the underlying contributors to the biological ageing process and point toward promising intervention targets to slow the biological ageing rate and promote healthy longevity.

## Data Availability

This study is based on publicly available data. The genetic association data of the 18 modifiable factors are available in Table 1. The GWAS summary statistics for GrimAgeAccel and PhenoAgeAccel are available at https://datashare.is.ed.ac.uk/handle/10283/3645.

## List of abbreviations

BF%: Body fat percentage
BMI: Body mass index
CI: Confidence interval
CpG: Cytosine-phosphate-guanine
CRP: C-reactive protein
DBP: Diastolic blood pressure
FDR: False discovery rate
GrimAgeAccel: GrimAge acceleration
GWAS: Genome-wide association study
HDL: High-density lipoprotein
IV: Instrumental variable
IVW: Inverse variance weighted
LDL: Low-density lipoprotein
MVPA: Moderate-to vigorous physical activity
MR: Mendelian randomization
MVMR: Multivariable Mendelian randomization
PhenoAgeAccel: PhenoAge acceleration
PRESSO: Pleiotropy residual sum and outlier
SBP: Systolic blood pressure
SD: Standard deviation
SE: Standard errors
SNP: Single nucleotide polymorphism
WM: Weighted median

## Ethics committee approval

This study is based on publicly available summarized data. Ethical approval and informed consent had been obtained in all original studies.

## Availability of data and materials

The genetic association data of the 18 modifiable factors are available in Supplementary Table 3. The GWAS summary statistics for GrimAgeAccel and PhenoAgeAccel are available at https://datashare.is.ed.ac.uk/handle/10283/3645. The analytical script of the MR analyses conducted in this study is available via the GitHub repository of the “TwoSampleMR” R package (https://github.com/MRCIEU/TwoSampleMR/).

## Authors’ contributions

LK and TW contributed to the conception and design of the study. LK and CY contributed to statistical analysis. LK contributed to drafting of the manuscript. TW guaranteed this work and take responsibility for the integrity of the data. All authors contributed to acquisition or interpretation of data, critical revision of the manuscript for important intellectual content, and final approval of the version to be published.

## Acknowledgements

We gratefully acknowledge the investigators and participants of all genome-wide association studies from which we used data.

