## Supplementary Table 1-5 for "Causal associations of education, lifestyle behaviors, and cardiometabolic traits with epigenetic age acceleration: a Mendelian randomization study"

### Supplementary files

#### Table of contents


| | |
|---|---|
| **Supplementary Table 1.** STROBE-MR checklist of the study | ……….……..2 |
| **Supplementary Table 2.** Detail definitions of exposures and outcomes in the MR study | ……………..…..5 |
| **Supplementary Table 3.** Data sources of IVs for the 18 modifiable factors used in the MR study | ……………..…..9 |
| **Supplementary Table 4.** Test for heterogeneity and pleiotropy in associations between 18 modifiable factors and GrimAgeAccel | ………..……11 |
| **Supplementary Table 5.** Test for heterogeneity and pleiotropy in associations between 18 modifiable factors and PhenoAgeAccel | ………..……12 |

**Supplementary Table 1. STROBE-MR checklist of the study[1]**

| Section | Item No. | Checklist item | Page No. |
|---|---|---|---|
| **Title and abstract** | 1 | Indicate Mendelian randomization (MR) as the study's design in the title and/or the abstract if that is a main purpose of the study | 1-5 |
| **Introduction** | | | |
| Background | 2 | Explain the scientific background and rationale for the reported study. What is the exposure? Is a potential causal relationship between exposure and outcome plausible? Justify why MR is a helpful method to address the study question | 6 |
| Objectives | 3 | State specific objectives clearly, including pre-specified causal hypotheses (if any). State that MR is a method that, under specific assumptions, intends to estimate causal effects | 6 |
| **Methods** | | | |
| Study design and data sources | 4 | Present key elements of the study design early in the article. Consider including a table listing sources of data for all phases of the study. For each data source contributing to the analysis, describe the following: | |
| | | a) Setting: Describe the study design and the underlying population, if possible. Describe the setting, locations, and relevant dates, including periods of recruitment, exposure, follow-up, and data collection, when available. | 7 |
| | | b) Participants: Give the eligibility criteria, and the sources and methods of selection of participants. Report the sample size, and whether any power or sample size calculations were carried out prior to the main analysis | 7, ST 3 |
| | | c) Describe measurement, quality control and selection of genetic variants | 7-8 |
| | | d) For each exposure, outcome, and other relevant variables, describe methods of assessment and diagnostic criteria for diseases | 7-8, ST 2 |
| | | e) Provide details of ethics committee approval and participant informed consent, if relevant | 7 |
| Assumptions | 5 | Explicitly state the three core IV assumptions for the main analysis (relevance, independence and exclusion restriction) as well assumptions for any additional or sensitivity analysis | 7, Figure 1 |
| Statistical methods: main analysis | 6 | Describe statistical methods and statistics used | |
| | | a) Describe how quantitative variables were handled in the analyses (i.e., scale, units, model) | 7, ST 3 |
| | | b) Describe how genetic variants were handled in the analyses and, if applicable, how their weights were selected | 7 |
| | | c) Describe the MR estimator (e.g. two-stage least squares, Wald ratio) and related statistics. Detail the included covariates and, in case of two-sample MR, whether the same covariate set was used for adjustment in the two samples | 9 |
| | | d) Explain how missing data were addressed | N/A |
| | | e) If applicable, indicate how multiple testing was addressed | 9 |
| Assessment of assumptions | 7 | Describe any methods or prior knowledge used to assess the assumptions or justify their validity | 9-10 |
| Sensitivity analyses and additional analyses | 8 | Describe any sensitivity analyses or additional analyses performed (e.g. comparison of effect estimates from different approaches, independent replication, bias | 9-10 |

| | | | |
|---|---|---|---|
| | | analytic techniques, validation of instruments, simulations) | |
| Software and pre-registration | 9 | a) Name statistical software and package(s), including version and settings used | 10 |
| | | b) State whether the study protocol and details were pre-registered (as well as when and where) | N/A |
| **Results** | | | |
| Descriptive data | 10 | a) Report the numbers of individuals at each stage of included studies and reasons for exclusion. Consider use of a flow diagram | 8, ST 3 |
| | | b) Report summary statistics for phenotypic exposure(s), outcome(s), and other relevant variables (e.g. means, SDs, proportions) | 7-8, ST 3 |
| | | c) If the data sources include meta-analyses of previous studies, provide the assessments of heterogeneity across these studies | N/A |
| | | d) For two-sample MR:<br>i. Provide justification of the similarity of the genetic variant-exposure associations between the exposure and outcome samples<br>ii. Provide information on the number of individuals who overlap between the exposure and outcome studies | 8 |
| Main results | 11 | a) Report the associations between genetic variant and exposure, and between genetic variant and outcome, preferably on an interpretable scale | 10-13, Table 1 and 2 |
| | | b) Report MR estimates of the relationship between exposure and outcome, and the measures of uncertainty from the MR analysis, on an interpretable scale, such as odds ratio or relative risk per SD difference | 10-13, Table 1 and 2 |
| | | c) If relevant, consider translating estimates of relative risk into absolute risk for a meaningful time period | NA |
| | | d) Consider plots to visualize results (e.g. forest plot, scatterplot of associations between genetic variants and outcome versus between genetic variants and exposure) | Figure 2 and 3 |
| Assessment of assumptions | 12 | a) Report the assessment of the validity of the assumptions | 11-13 |
| | | b) Report any additional statistics (e.g., assessments of heterogeneity across genetic variants, such as I2, Q statistic or E-value) | 11-13, ST 4-5 |
| Sensitivity analyses and additional analyses | 13 | a) Report any sensitivity analyses to assess the robustness of the main results to violations of the assumptions | 11-13, Table 1 and 2 |
| | | b) Report results from other sensitivity analyses or additional analyses | 11-13, Table 1 and 2 |
| | | c) Report any assessment of direction of causal relationship (e.g., bidirectional MR) | NA |
| | | d) When relevant, report and compare with estimates from non-MR analyses | 13-16 |
| | | e) Consider additional plots to visualize results (e.g., leave-one-out analyses) | NA |
| **Discussion** | | | |
| Key results | 14 | Summarize key results with reference to study objectives | 13 |
| Limitations | 15 | Discuss limitations of the study, taking into account the validity of the IV assumptions, other sources of potential bias, and imprecision. Discuss both direction and magnitude of any potential bias and any efforts to address them | 16-17 |

| | | | |
|---|---|---|---|
| Interpretation | 16 | a) Meaning: Give a cautious overall interpretation of results in the context of their limitations in comparison with other studies | 14-17 |
| | | b) Mechanism: Discuss underlying biological mechanisms that could drive a potential causal relationship between the investigated exposure and the outcome, and whether the gene-environment equivalence assumption is reasonable. Use causal language carefully, clarifying that IV estimates may provide causal effects only under certain assumptions | 14-17 |
| | | c) Clinical relevance: Discuss whether the results have clinical or public policy relevance, and to what extent they inform effect sizes of possible interventions | 14-17 |
| Generalizability | 17 | Discuss the generalizability of the study results (a) to other populations, (b) across other exposure periods/timings, and (c) across other levels of exposure | 17 |
| **Other information** | | | |
| Funding | 18 | Describe sources of funding and the role of funders in the present study and, if applicable, sources of funding for the databases and original study or studies on which the present study is based | 19 |
| Data and data sharing | 19 | Provide the data used to perform all analyses or report where and how the data can be accessed, and reference these sources in the article. Provide the statistical code needed to reproduce the results in the article, or report whether the code is publicly accessible and if so, where | 19 |
| Conflicts of Interest | 20 | All authors should declare all potential conflicts of interest | 19 |

Abbreviations: ST=supplementary table.

**Supplementary Table 2. Detail definitions of exposures and outcomes in the MR study**

| Phenotype | Definition | References |
|---|---|---|
| **Exposure** | | |
| **Socioeconomic factor** | | |
| Educational attainment | Educational attainment was measured as the number of years of schooling that individuals completed (EduYears). The EduYears phenotype was categorized according to the International Standard Classification of Education (ISCED) 2011, converted to US years of schooling and standardized, with each unit (SD) representing 4.2 years of schooling. | 1 |
| **Lifestyle factor** | | |
| Smoking initiation | Smoking initiation was defined as a binary phenotype indicating whether an individual had ever smoked regularly. Participants who reported ever being a regular smoker in their life (current or former) were defined as smokers; the remaining participants were defined as non-smokers. | 2 |
| Alcohol intake | Alcohol intake was measured by drinks per week. Drinks per week was defined as the average number of drinks a participant reported drinking each week, aggregated across all types of alcohol.<br>If a study recorded binned response ranges (e.g., 1-4 drinks per week, 5-10 drinks per week), then used the midpoint of the range. This phenotype was left-anchored at 1 and log-transformed prior to analysis, in order to prevent outliers from having undue leverage on analyses. | 2 |
| Coffee consumption | Coffee intake was collected using a 24-hour recall questionnaire (Oxford WebQ) in a subset of UK Biobank participants. The mean intake from participants who completed at least two dietary recalls was used in the genome-wide association study (GWAS) study. | 3 |
| Daytime napping | Self-reported daytime napping was ascertained in the UK Biobank using the question “Do you have a nap during the day?” with responses options of “Never/rarely”, “Sometimes”, “Usually” and “Prefer not to answer”. Prefer not to answer responses were set to missing. Responses were treated as a continuous variable in the GWAS. | 4 |
| Sleep duration | Study participants were asked “About how many hours sleep do you get in every 24 h”, with responses in hour increments. Sleep duration was treated as a continuous variable. Extreme responses of less than 3 h or more than 18 h were excluded and “Do not know or Prefer not to answer” responses were set to missing. Participants who self-reported any sleep medication were excluded. | 5 |
| MVPA | Moderate-to-vigorous physical activity (MVPA) was calculated by taking the sum of total minutes/week of MPA multiplied by four and the total number of VPA minutes/week multiplied by eight, corresponding to their metabolic equivalents. | 6 |
| **Cardiometabolic factor** | | |
| Adiposity-related traits | GWAS data for body mass index (BMI) and waist circumference were from the Genetic Investigation of Anthropometric Traits (GIANT) consortium. BMI was calculated by dividing weight (kg) by height squared (m2). Waist circumferences were measured or self-reported.<br>GWAS data for body fat percentage was from the UK Biobank, where body fat percentage was measured using the Tanita BC418MA body composition analyzer. Body fat percentage (BF%) was estimated by impedance measurement.<br>Childhood obesity cases were defined as participants having | 7-10 |

| | | |
|---|---|---|
| | ≥95th percentile of BMI for age, whereas childhood normal weight controls were defined as having a BMI < 50th percentile. | |
| Type 2 diabetes | GWAS data for type 2 diabetes without adjustment for BMI was from the DIAbetes Genetics Replication And Meta-analysis (DIAGRAM) consortium. In the original GWAS, type 2 diabetes was defined by diagnostic fasting glucose, casual glucose, 2 h plasma glucose or HbA1c levels; use of glucose-lowering medication (by Anatomical Therapeutic Chemical code or self-report); or type 2 diabetes history from electronic medical records, self-report and varying combinations of each, depending on the contributing cohort. | 11 |
| Lipids traits | Data for low-density lipoprotein (LDL) cholesterol, triglycerides, and high-density lipoprotein (HDL) cholesterol were from the Million Veteran Program (MVP) and the Global Lipids Genetics Consortium (GLGR).<br>Following extraction of prevalent laboratory measurements from the electronic health record, lipid data were evaluated for spurious values (<0 mg/dL), histograms for each lipid trait were inspected for normality, and extreme outliers (>400 mg/dL, >1000 mg/dL, >500 mg/dL, and >150 mg/dL for LDL-C, triglycerides, and HDL-C, respectively were excluded. For each phenotype: maximum LDL-C, natural log transformed maximum triglycerides, and minimum HDL-C, residuals were obtained after regressing on age, sex, and 10 principal components within each ethnic group. Residuals were subsequently inverse normal transformed for association analysis. To minimize confounding from statins and variable adherence, maximum/minimum values were used. | 12 |
| Blood pressure | GWAS data for systolic blood pressure (SBP) and diastolic blood pressure (DBP) were from the UK Biobank and ICBP. SBP and DBP values were the mean of two automated or two manual blood pressure measurements.<br>The UK Biobank comprised observational and genotyping data of 502,519 people aged between 40 and 69 years. Following informed consent participants completed a standardized questionnaire on life course exposures, medical history and treatments and underwent a standardized portfolio of phenotypic tests including two blood pressure measurements taken seated after two minutes rest using an appropriate cuff and an Omron HEM-7015IT digital blood pressure monitor. A manual sphygmometer was used if the standard automated device could not be employed.<br>The ICBP GWAS dataset consisted of 299,024 individuals of European ancestry from a total of 77 cohorts, and the assessment details of SBP and DBP varied among cohorts. Here we listed the definitions of SBP and DBP in two cohorts as examples, for more details please refer to the Supplementary Table 1b of the paper by Evangelou E, et al. (13).<br>1) In the AGES study, participants came in a fasting state to the clinic. The supine blood pressure was measured twice by a nurse using a mercury sphygmomanometer after a 5-min rest.<br>2) In the ARIC study, blood pressure was measured using a standardized Hawksley random-zero mercury column sphygmomanometer with participants in a sitting position after a resting period of 5 minutes. The size of the cuff was chosen according to the arm circumference. Three sequential | 13 |

| | | |
|---|---|---|
| | recordings for SBP and DBP were obtained; the mean of the last two measurements was used in this analysis, discarding the first reading. Outliers of >4SD were discarded. | |
| CRP | The data for C-reactive protein (CRP) was from the UK Biobank (Data-Field 30710). C-reactive protein (mg/L) was measured by immunoturbidimetric - high sensitivity analysis on a Beckman Coulter AU5800. | 14 |
| **Outcome** | | |
| Epigenetic age acceleration metrics (GrimAgeAccel and PhenoAgeAccel) | DNAm GrimAge (in units of years), incorporates data from 1,030 CpGs based on the seven plasma proteins (i.e. cystatin C, leptin, tissue inhibitor metalloproteinases 1, adrenomedullin, beta-2-microglobulin, growth differentiation factor 15, and plasminogen activation inhibitor 1) and smoking pack-years. The GrimAge acceleration (GrimAgeAccel) is the raw residuals in the linear regression models with DNAm GrimAge regressed on chronological age and sex.<br>DNAm PhenoAge (in units of years) algorithm involves 513 CpGs based on chronological age and nine clinical biomarkers (i.e. albumin, creatinine, serum glucose, C-reactive protein, lymphocyte percentage, mean corpuscular volume, red cell distribution width, alkaline phosphatase and leukocyte count). The PhenoAge acceleration (PhenoAgeAccel) is the residuals of the regression models that regress DNAm PhenoAge on chronological age.<br>The meta-analysis samples with DNA methylation data were from 28 cohorts of 34,710 European participants.<br>Age-adjusted DNA methylation-based estimates of GrimAge and PhenoAge were calculated using the Horvath epigenetic age calculator software (https://dnamage.genetics.ucla.edu/) or standalone scripts. The following outputs were assessed: GrimAge acceleration—"GrimAgeAccel" and PhenoAge acceleration—"PhenoAgeAccel". Outlier samples with clock methylation estimates of +/−5 standard deviations from the mean were excluded from further analysis. | 15 |

Abbreviations: BF%=Body fat percentage; BMI=body mass index; CRP=C-reactive protein; DBP=diastolic blood pressure; DIAGRAM=DIAbetes Genetics Replication And Meta-analysis; GrimAgeAccel=GrimAge acceleration; GIANT=Genetic Investigation of Anthropometric Traits; GLGC=Global Lipids Genetics Consortium; GWAS=genome-wide association study; HDL=high-density lipoprotein; LDL=low-density lipoprotein; MVP=Million Veteran Program; MVPA=moderate-to vigorous physical activity; PhenoAgeAccel=PhenoAge acceleration; SBP=systolic blood pressure; SD=standard deviation; VPA=vigorous physical activity.

**Supplementary Table 3. Data sources of IVs for the 18 modifiable factors used in the MR study**

| Modifiable factor | PMID/ GWAS ID | Sample size | Ancestry | No. of SNPs[a] | Unit | P threshold[b] |
|---|---|---|---|---|---|---|
| **Socioeconomic factor** | | | | | | |
| Educational attainment[1] | 30038396 | 1131881 | European | 751 | 1-SD increase in years of schooling | P $<5\times10^{-8}$ |
| **Lifestyle behavior** | | | | | | |
| Smoking initiation[2] | 30643251 | 557337 cases and 674754 controls | European | 299 | log-odds (ever smoked regularly compared to never smoked) | P $<5\times10^{-8}$ |
| Alcohol intake[2] | 30643251 | 941280 | European | 80 | 1-SD increase in log-transformed alcoholic drinks/week | P $<5\times10^{-8}$ |
| Coffee consumption[3] | 31046077 | 375833 | European | 12 | 1% change | P $<5\times10^{-8}$ |
| Sleep | | | | | | |
| Daytime napping[4] | 33568662 | 452633 | European | 115 | 1 unit increase in napping category (responses “never, sometimes or usually napping” were treated as continuous variable) | P $<5\times10^{-8}$ |
| Sleep duration[5] | 30846698 | 446118 | European | 77 | 1 hour/day | P $<5\times10^{-8}$ |
| MVPA[6] | 29899525 | 377234 | European | 6 | 1-SD increase in MET-minutes/week of MVPA | P $<5\times10^{-9}$ |
| **Cardiometabolic trait** | | | | | | |
| Adiposity | | | | | | |
| BMI[7] | 30124842 | 681275 | European | 941 | 1-SD increase in body mass index | P $<5\times10^{-8}$ |
| Waist circumference[8] | 25673412 | 224459 | European | 44 | 1-SD increase in waist circumference | P $<5\times10^{-8}$ |
| BF% | ukb-b-8909 | 454633 | European | 641 | 1-SD increase in body fat percentage | P $<5\times10^{-8}$ |

| | | | | | | |
|---|---|---|---|---|---|---|
| Childhood obesity[9] | 22484627 | 5530 cases and 8318 controls | European | 5 | log-odds | P $<5\times10^{-8}$ |
| Type 2 diabetes[10] | 30297969 | 71124 cases and 824006 controls | European | 232 | log-odds | P $<5\times10^{-8}$ |
| Lipids[11] | | | | | | |
| LDL cholesterol | 30275531 | > 600000 | mix | 145 | 1-SD increase in LDL cholesterol | P $<5\times10^{-8}$ |
| HDL cholesterol | 30275531 | > 600000 | mix | 222 | 1-SD increase in HDL cholesterol | P $<5\times10^{-8}$ |
| Triglycerides | 30275531 | > 600000 | mix | 172 | 1-SD increase in triglycerides | P $<5\times10^{-8}$ |
| Blood pressure[12] | | | | | | |
| SBP | 30224653 | >1 million | European | 222 | 1 mmHg | P $<5\times10^{-8}$ |
| DBP | 30224653 | >1 million | European | 264 | 1 mmHg | P $<5\times10^{-8}$ |
| CRP[13] | 31900758 | 418642 | European | 299 | 1-SD increase in serum CRP levels | P $<5\times10^{-8}$ |

[a]SNPs used in the present MR analysis.

[b]P threshold represents genome-wide significance threshold of genetic instruments.

Abbreviations: BF%=body fat percentage; BMI=body mass index; CRP=C-reactive protein; DBP=diastolic blood pressure; GWAS=genome-wide association study; HDL=high-density lipoprotein; IVs=instrumental variables; LDL=low-density lipoprotein; MVPA=moderate-to vigorous physical activity; MR=Mendelian randomization; No=number; SBP=systolic blood pressure; SD=standard deviation; SNP=single nucleotide polymorphism.

**Supplementary Table 4. Test for heterogeneity and pleiotropy in associations between 18 modifiable factors and GrimAgeAccel**

| Modifiable factor | Pleiotropy test | | Heterogeneity test | |
|---|---|---|---|---|
| | **Egger intercept (SE)** | **$P_{intercept}$** | **Q statistic** | **$P_h$** |
| **Socioeconomic factor** | | | | |
| Educational attainment | -0.004 (0.005) | 0.471 | 865.51 | 0.002 |
| **Lifestyle factor** | | | | |
| Smoking initiation | -0.0004 (0.009) | 0.959 | 324.13 | 0.143 |
| Alcohol intake | 0.006 (0.009) | 0.518 | 105.49 | 0.025 |
| Coffee consumption | 0.030 (0.024) | 0.249 | 16.07 | 0.138 |
| Sleep | | | | |
| Daytime napping | 0.004 (0.012) | 0.740 | 122.38 | 0.279 |
| Sleep duration | -0.004 (0.017) | 0.820 | 87.22 | 0.178 |
| MVPA | 0.138 (0.083) | 0.173 | 3.57 | 0.613 |
| **Cardiometabolic factor** | | | | |
| Adiposity | | | | |
| BMI | -0.003 (0.003) | 0.361 | 1032.48 | 0.019 |
| Waist circumference | -0.003 (0.015) | 0.835 | 49.35 | 0.234 |
| BF% | -0.001 (0.005) | 0.804 | 756.87 | 0.001 |
| Childhood obesity | -0.115 (0.107) | 0.362 | 3.28 | 0.512 |
| Type 2 diabetes | 0.004 (0.006) | 0.523 | 280.16 | 0.015 |
| Lipids | | | | |
| LDL cholesterol | -0.006 (0.005) | 0.290 | 203.53 | 8.06E-04 |
| HDL cholesterol | -0.004 (0.005) | 0.431 | 311.31 | 7.08E-05 |
| Triglycerides | 0.004 (0.005) | 0.423 | 175.27 | 0.416 |
| Blood pressure | | | | |
| SBP | -0.003 (0.010) | 0.744 | 248.87 | 0.096 |
| DBP | -0.009 (0.008) | 0.259 | 295.77 | 0.080 |
| CRP | 0.013 (0.003) | 7.23E-05 | 348.43 | 0.023 |

GrimAgeAccel represents epigenetic-age acceleration obtained using the GrimAge clock.
Abbreviations: BF%=body fat percentage; BMI=body mass index; CRP=C-reactive protein; DBP=diastolic blood pressure; HDL=high-density lipoprotein; LDL=low-density lipoprotein; MVPA=moderate-to vigorous physical activity; $P_{intercept}$=P of intercept; $P_h$=P of heterogeneity test; SBP=systolic blood pressure.

**Supplementary Table 5. Test for heterogeneity and pleiotropy in associations between 18 modifiable factors and PhenoAgeAccel**

| Modifiable factor | Pleiotropy test | | Heterogeneity test | |
|---|---|---|---|---|
| | **Egger intercept (SE)** | **$P_{intercept}$** | **Q statistic** | **$P_h$** |
| **Socioeconomic factor** | | | | |
| Educational attainment | -0.004 (0.006) | 0.512 | 838.24 | 0.013 |
| **Lifestyle factor** | | | | |
| Smoking initiation | -0.014 (0.011) | 0.220 | 356.87 | 0.011 |
| Alcohol intake | -0.015 (0.012) | 0.210 | 96.19 | 0.091 |
| Coffee consumption | 0.019 (0.036) | 0.616 | 20.35 | 0.041 |
| Sleep | | | | |
| Daytime napping | -0.0004 (0.016) | 0.982 | 152.69 | 0.009 |
| Sleep duration | -0.056 (0.022) | 0.013 | 98.52 | 0.042 |
| MVPA | 0.045 (0.104) | 0.691 | 1.63 | 0.898 |
| **Cardiometabolic factor** | | | | |
| Adiposity | | | | |
| BMI | 0.001 (0.004) | 0.788 | 1070.53 | 0.002 |
| Waist circumference | 0.003 (0.022) | 0.873 | 65.66 | 0.015 |
| BF% | -0.013 (0.006) | 0.050 | 761.87 | 0.001 |
| Childhood obesity | 0.017 (0.139) | 0.909 | 3.12 | 0.538 |
| Type 2 diabetes | 0.005 (0.007) | 0.507 | 273.45 | 0.029 |
| Lipids | | | | |
| LDL cholesterol | -0.006 (0.007) | 0.393 | 207.96 | 3.90E-04 |
| HDL cholesterol | -0.009 (0.006) | 0.131 | 255.49 | 0.061 |
| Triglycerides | 0.005 (0.006) | 0.375 | 192.43 | 0.137 |
| Blood pressure | | | | |
| SBP | 0.010 (0.013) | 0.478 | 273.03 | 0.010 |
| DBP | -0.015 (0.011) | 0.165 | 293.81 | 0.093 |
| CRP | 0.015 (0.004) | 4.56E-04 | 368.54 | 0.003 |

PhenoAgeAccel represents epigenetic-age acceleration obtained using the PhenoAge clock. Abbreviations: BF%=body fat percentage; BMI=body mass index; CRP=C-reactive protein; DBP=diastolic blood pressure; HDL=high-density lipoprotein; LDL=low-density lipoprotein; MVPA=moderate-to vigorous physical activity; $P_{intercept}$=P of intercept; $P_h$=P of heterogeneity test; SBP=systolic blood pressure.
